# Medical students’ perceptions and motivations in time of COVID-19 pandemic

**DOI:** 10.1101/2020.05.28.20115956

**Authors:** Patricia Tempski, Fernanda M. Arantes-Costa, Renata Kobayasi, Marina A.M. Siqueira, Matheus B. Torsani, Bianca Q.R.C. Amaro, Maria Eduarda FM Nascimento, Saulo L Siqueira, Itamar S. Santos, Milton A. Martins

## Abstract

**Background:** There has been a rapid increase in the number of cases of COVID-19 in Latin America, Africa and Asia, in many countries that have an insufficient number of physicians and other health care personnel, and the need for the inclusion of medical students as part of the health teams is a very important issue. It has been recommended that medical students work as volunteers, have appropriate training, do not undertake any activity beyond their level of competence, have continuous supervision and adequate personal protective equipment. However, motivation of medical students must be evaluated in order to make volunteering a more evidence-based initiative. The aim of our study was to evaluate motivation of medical students to be part of the heath team tohelp in the COVID-19 pandemic.

**Methods and Findings:** We developed a questionnaire specifically to evaluate medical student’s perceptions about participating in care of patients with suspected infection due to coronavirus during the COVID-19 pandemic. The questionnaire had two parts: a) individual characteristics, year and geographic location of medical school; b) twenty-eight statements responded on a 5-point Likert scale (totally agree, agree, neither agree nor disagree, disagree and totally disagree). To develop the questionnaire, we performed consensus meetings of a group of faculty and medical students. The questionnaire was sent to student organizations of 257 medical schools in Brazil and answered by 10,433 students. We used multinomial logistic regression models to analyse the data.

Statements with greater odds ratios for participation of medical students in COVID-19 pandemic were related to sense of purpose or duty (“It is the duty of the medical student to put himself at the service of the population in the pandemic”), altruism (“I am willing to take risks by participating in practical in the context of pandemic”), perception of good performance and professional identity (“I will be a better health professional for having experienced the pandemic”). Males had higher odds ratios than females (1.36 [95% CI: 1.24 – 1.49] to 1.68 [95% CI: 1.47 – 1.91]).

**Conclusions:** Medical students are motivated by sense of purpose or duty, altruism, perception of good performance and values of professionalism more than their interest in learning. These results have implications in the developing of programs of volunteering and in the design of health force policies in the present pandemic and in future health emergencies.

## Introduction

The coronavirus COVID-19 pandemic is the most important global health crisis of our time and the greatest challenge the health system has faced since World War Two. Since its emergence in Asia in 2019, the virus has spread to every continent, except Antarctica. Cases are rising daily in Europe, North America, and, in the last weeks, also in Latin America and Africa. [1,2]

COVID-19 pandemic resulted in a disruption of undergraduate medical education. In many countries, medical education faculty have quickly transitioned the first-years curriculum to on-line activities in response to the need of social isolation to flatten the curve of new cases of COVID-19. [3] In addition, in the final years of medical schools, in many countries, clerkships were severely affected by the rapid changes in hospitals due to the need of care of an increasing number of COVID-19 patients and medical students were advised to stay at home, considering the potential risk of medical students spreading COVID-19 infection in health care settings and the shortage of Personal Protective Equipment (PPE). [3,4]

However, the role of medical students in the COVID-19 pandemic is changing rapidly, due to the shortage of health professionals in many cities, even in developed countries. Both the Medical Schools Council (MSC) of United Kingdom and the American Association of Medical Colleges (AAMC) of the United States have published guidelines for the participation of medical students in the global effort to give the best care to patients with COVID-19. [5,6] Both associations recommend that medical students work as volunteers, have appropriate training, do not undertake any activity beyond their level of competence, have continuous supervision and adequate PPE. [5,6]

In the last weeks, there has been a huge increase in the number of cases in countries that have an insufficient number of physicians and other health care personnel, and it is possible to anticipate the need for the inclusion of medical students as part of the health teams. [2,7]

In many countries thousands of medical students have volunteered their services to support the fight against the coronavirus pandemic. [8,9] Motivation is pivotal to volunteering and must be evaluated in order to make volunteering a more evidence-based The aim of our study was to evaluate motivation of medical students to be part of the heath team to help in the COVID-19 pandemic. It was performed in a developing country, Brazil, in the first week of the increase in cases of COVID-19 in Brazil, including 10,433 medical students. initiative.

## Materials and Methods

We developed a questionnaire specifically to evaluate medical student’s perceptions about participating in care of patients with suspected infection due to coronavirus during the COVID-19 pandemic. The questionnaire had two parts: a) individual characteristics, year and geographic location of medical school; b) twenty-eight statements responded on a 5-point Likert scale (totally agree, agree, neither agree nor disagree, disagree and totally disagree). To develop the questionnaire, we performed consensus meetings of a group of faculty and medical students.

The questionnaire was developed on a Google Form Survey Administration App. With the help of the Brazilian Section of the International Federation of Medical Students Associations (IFMSA), the questionnaire was sent to medical student organizations of medical schools of all regions of Brazil, using Instagram and WhatsApp social media. In the first page of the questionnaire, the purpose of the study was explained, and the student had to agree with the objectives of the study before filling the questionnaire.

The survey was performed between 20^th^ and 22^th^ of March, since after these three days of survey we had received more than 10,000 answers.

In this period, there were only a small number of patients with the diagnosis of COVID-19 disease in Brazil; 1,546 confirmed cases and 25 deaths due to COVID-19, according to the Ministry of Health of Brazil. [10]

In Brazil, undergraduate medical course is a six-year program, with four years of basic and clinical sciences and two years of clerkships (internship) when medical students have responsibilities with the direct care of patients under the supervision of faculty or preceptors of the National Health System.

There are 341 medical schools in Brazil with about 35,288 first year medical students. [11] The questionnaire was sent to the student organizations of 257 medical schools (75.4% of Brazilian medical schools).

### Study variables

Participants informed their agreement or not with two statements regarding their view about the role medical students should have during the COVID-19 pandemic. These questions were (S8) Medical internship students must participate in health care assistance during pandemic; and (S9) All students regardless their year during the medical school, must participate in health care assistance during pandemic. Participants were classified according to their agreement with statements S8 and S9 as: (1) All students should participate (agree with S9); (2) Only students in internship should participate (not agree nor disagree / disagree with S9 and agree with S8); and (3) No students should participate (not agree nor disagree / disagree with S9, not agree nor disagree / disagree with S8).

For the analysis, we combined the responses “completely agree” with “agree” for each statement.

### Statistical analysis

Continuous variables are expressed as means ± standard deviations. Categorical variables are expressed as absolute counts and proportions and compared across groups using chi-squared tests.

We used multinomial logistic regression models to study the association between student’s characteristics and perceptions according to their opinion about student’s participation in the healthcare of COVID-19 pandemic. Models are presented (1) crude and (2) adjusted for sex, year of medical school and geographic region of the country. Analyses were performed using IBM Corp. Released 2013, SPSS Statistics for Windows, Version 22.0. Armonk, NY and R software version 3.4.4 (Vienna, Austria).

## Results

Table 1 shows the number of medical students from the five geographic regions of the country that answered the questionnaire, and the relationship between the number of participants and the number of vacancies in each region of the country. There was a similar proportion of respondents/vacancies, showing that the sample was homogenously distributed across the country.

**Table 1:**
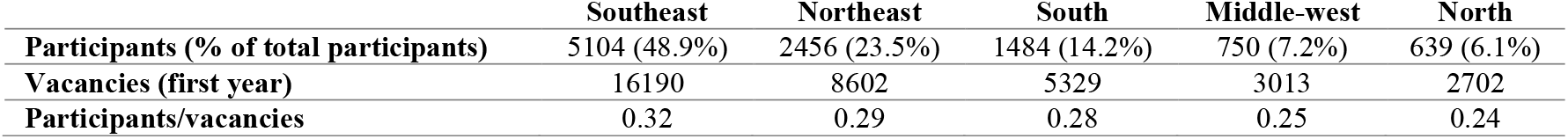
Participants distribution according to geographic region of Brazil.

|  | Southeast | Northeast | South | Middle-west | North |
| --- | --- | --- | --- | --- | --- |
| **Participants (% of total participants)** | 5104 (48.9%) | 2456 (23.5%) | 1484 (14.2%) | 750 (7.2%) | 639 (6.1%) |
| **Vacancies (first year)** | 16190 | 8602 | 5329 | 3013 | 2702 |
| **Participants/vacancies** | 0.32 | 0.29 | 0.28 | 0.25 | 0.24 |

Table 2 shows the number (and percentage) of medical students that answered “completely agree” or “agree” for each one of the 28 statements of the questionnaire. Students were divided considering year of medical course and sex. Supplemental Table 1 shows the answers to the questionnaire for the hole group of medical students.

**Table 2:** Students perceptions according to year of medical course and sex during the COVID-19 pandemic –number of students that responded completely agree or agree (% of total)

|  | Year of graduation |  |  | Gender |  |
| --- | --- | --- | --- | --- | --- |
|  | 1<sup>st</sup>/2<sup>nd</sup> | 3<sup>rd</sup>/4<sup>th</sup> | 5<sup>th</sup>/6<sup>th</sup> | Female | Male |
| S1 I feel prepared to identify a suspected case*# | 2252 (49.3) | 2262 (66.1) | 1879 (76.9) | 4277 (58.9) | 2116 (66.8) |
| S2 I can identify signs of severity in a patient*# | 2721 (59.6) | 2760 (80.7) | 2169 (88.7) | 5182 (71.3) | 2468 (78.0) |
| S3 I know how to guide for preventive measures# | 4316 (94.5) | 3310 (96.8) | 2341 (95.7) | 6962 (94.9) | 3005 (95.8) |
| S4 I know how to guide for therapeutic measures*# | 984 (21.5) | 1037 (30.3) | 1239 (50.7) | 2156 (29.7) | 1104 (34.9) |
| S5 I know how to use personal protection equipment (PFE)# | 3615 (79.2) | 2773 (81.1) | 1867 (76.4) | 5749 (79.1) | 2506 (79.2) |
| S6 I am able to participate in the care of patients who seek health care*# | 849 (18.6) | 1230 (36.0) | 1370 (56.0) | 2206 (30.4) | 1243 (39.3) |
| S7 I feel able to communicate the diagnosis for COVID-19 infection*# | 1153 (25.2) | 1435 (41.9) | 1423 (58.2) | 2489 (34.3) | 1522 (48.1) |
| S8 Medical internship students must participate in health care assistance during pandemic*# | 3054 (66.9) | 2011 (58.8) | 1225 (50.1) | 4206 (57.9) | 2084 (65.8) |
| S9 All students regardless their year during the course, must participate in health care assistance during pandemic*# | 830 (18.2) | 478 (14.0) | 90 (3.7) | 888 (12.2) | 510 (16.1) |
| S10 It is the duty of the medical student to put himself at the service of the population in the pandemic*# | 2190 (48.0) | 1433 (41.9) | 853 (34.9) | 3005 (41.4) | 1471 (46.5) |
| S11 I feel insecure regarding the future*# | 2786 (61.0) | 2217 (64.8) | 1766 (72.2) | 5068 (69.7) | 1701 (53.7) |
| S12 I am afraid of contaminating myself*# | 2705 (59.2) | 2055 (60.1) | 1673 (68.4) | 4695 (64.6) | 1738 (54.9) |
| S13 Medical schools must suspend their academic activities during the first to fourth year# | 3365 (73.7) | 2563 (74.9) | 2197 (89.9) | 5670 (78.0) | 2455 (77.5) |
| S14 Medical schools must suspend their academic activities during internship*# | 1175 (25.7) | 1130 (33.0) | 1061 (43.4) | 2435 (33.5) | 931 (29.4) |
| S15 Distance learning must be implemented in the suspension of academic activities*# | 2322 (50.8) | 2186 (63.9) | 1634 (66.8) | 4443 (61.1) | 1699 (53.7) |
| S16 I prefer to delay my training to fully replace academic activities than to participate in distance learning activities*# | 1813 (39.7) | 991 (29.0) | 715 (29.2) | 2304 (31.7) | 1215 (38.4) |
| S17 After pandemic, academic activities must be fully restored *# | 3017 (66.1) | 1917 (56.0) | 1178 (48.2) | 4125 (56.8) | 1987 (62.8) |
| S18 After pandemic, only practical academic activities must be restored*# | 1699 (37.2) | 1665 (48.7) | 985 (40.3) | 3167 (43.6) | 1182 (37.3) |
| S19 I feel able to study my medical course contents at distance# | 1913 (41.9) | 1927 (56.3) | 1438 (58.8) | 3655 (50.3) | 1623 (51.3) |
| S20 I prefer to study theoretical content using distance learning methods*# | 1604 (35.1) | 1631 (47.7) | 1322 (54.1) | 3267 (45.0) | 1290 (40.7) |
| S21 My emotional state during the pandemic affects my learning*# | 1856 (40.6) | 1319 (38.6) | 1089 (44.5) | 3217 (44.3) | 1047 (33.1) |
| S22 I will be a better health professional for having experienced the pandemic*# | 2606 (57.0) | 1936 (56.6) | 1393 (57.0) | 4020 (55.3) | 1914 (60.5) |
| S23 I feel stressed in the hospital at the moment*# | 1168 (25.6) | 1215 (35.5) | 1226 (50.1) | 2675 (36.8) | 934 (29.5) |
| S24 The supervision I receive in my practices fields is good*# | 1874 (41.0) | 1820 (53.2) | 1187 (48.5) | 3324 (45.7) | 1557 (49.2) |
| S25 I have access to psychological support*# | 2201 (48.2) | 1431 (41.8) | 747 (30.6) | 2938 (40.4) | 1441 (45.5) |
| S26 I am proud of the way my institution responded to social and health demands in the face of the pandemic*# | 2653 (58.1) | 1905 (55.7) | 991 (40.5) | 3798 (52.3) | 1751 (55.3) |
| **S27 The role of the medical students during the pandemics time is irrelevant\*#** | 185 (4.1) | 204 (6.0) | 275 (11.2) | 463 (6.4) | 201 (6.3) |
| **S28 I am willing to take risks by participating in practical in the context of pandemic\*#** | 1967 (43.1) | 1581 (46.2) | 1282 (52.4) | 3144 (43.3) | 1686 (53.3) |

Chi-square test. gender\* $P < 0.05$
Chi-square test year of graduation # $P < 0.05$

Of the 10,433 students who completed the survey, 7,267 (69.7%) were females, and 2,445 (23.4%) were in internship years (interns). The mean age was 22.5 ± 3.9 years.

All students should participate in the response to the COVID-19 pandemic according to 1,398 (13.4%) participants. Only students in internship should participate according to 4,963 (47.6%), and no students should participate for 4,072 (39.0%) respondents.

Table 3 shows the participants’ characteristics and perceptions according to their view about the role of medical students during the COVID-19 pandemic. The statements which most participants agreed to were:

S3 “I know how to guide for preventive measures” (95.5%),

S5 “I know how to use personal protection equipment (PFE)” (79.1%), and

S13 “Medical schools must suspend their academic activities during the first to fourth year” (77.9%).

**Table 3.** Students characteristics and perceptions according to their view about the role of medical students during the COVID-19 pandemic. Students were divided into three groups according to their opinion about the participation of medical students in the care of patients with COVID-19. We show the number and percentage of medical students that answered totally agree or agree to each one of the statements.

|  | No students<br>should<br>participate<br>(N=4072) | Only students in<br>internship should<br>participate<br>(N=4963) | All students<br>should<br>participate<br>(N=1398) | Total<br>(N=10433) |
| --- | --- | --- | --- | --- |
| **Students' characteristics** |  |  |  |  |
| Age (mean ± SD) | 22.7 ± 3.7 | 22.4 ± 3.9 | 22.4 ± 4.2 | 22.5 ± 3.9 |
| Female sex (N (%)) | 3008 (73.9%) | 3371 (67.9%) | 888 (63.5%) | 7267 (69.7%) |
| Personal / family / friend diagnosis of COVID-19<br>(N (%)) | 308 (7.6%) | 334 (6.7%) | 85 (6.1%) | 727 (7.0%) |
| Year of medical school (N (%)) |  |  |  |  |
| First/second year (Basic sciences) | 1472 (36.1%) | 2265 (45.6%) | 830 (59.4%) | 4567 (43.8%) |
| Third/fourth (Clinical sciences) | 1391 (34.2%) | 1552 (31.3%) | 478 (34.2%) | 3421 (32.8%) |
| Fifth/sixth (Internship) | 1209 (29.7%) | 1146 (23.1%) | 90 (6.4%) | 2445 (23.4%) |
| **Students' perceptions** |  |  |  |  |
| S3. I know how to guide for preventive measures | 3805 (93.4%) | 4811 (96.9%) | 1351 (96.6%) | 9967 (95.5%) |
| S5. I know how to use personal protection<br>equipment (PFE) | 2988 (73.4%) | 4068 (82.0%) | 1199 (85.8%) | 8255 (79.1%) |
| S13. Medical schools must suspend their<br>academic activities during the first to fourth<br>year. | 3436 (84.4%) | 3925 (79.1%) | 764 (54.6%) | 8125 (77.9%) |
| S2. I can identify signs of severity in a patient | 2853 (70.1%) | 3701 (74.6%) | 1096 (78.4%) | 7650 (73.3%) |
| S11. I feel insecure regarding the future | 2835 (69.6%) | 3117 (62.8%) | 817 (58.4%) | 6769 (64.9%) |
| S12. I am afraid of contaminating myself | 2937 (72.1%) | 2829 (57.0%) | 667 (47.7%) | 6433 (61.7%) |
| S1. I feel prepared to identify a suspected case | 2270 (55.7%) | 3173 (63.9%) | 950 (68.0%) | 6393 (61.3%) |
| S15. Distance learning must be implemented in<br>the suspension of academic activities | 2446 (60.1%) | 2899 (58.4%) | 797 (57.0%) | 6142 (58.9%) |
| S17. After pandemic, academic activities must be<br>fully restored | 2262 (55.6%) | 2913 (58.7%) | 937 (67.0%) | 6112 (58.6%) |
| S22. I will be a better health professional for<br>having experienced the pandemic | 1807 (44.4%) | 3105 (62.6%) | 1022 (73.1%) | 5934 (56.9%) |
| S26. I am proud of the way my institution<br>responded to social and health demands in the<br>face of the pandemic | 1958 (48.1%) | 2758 (55.6%) | 833 (59.6%) | 5549 (53.2%) |
| S19. I feel able to study my medical course<br>contents at distance | 2040 (50.1%) | 2533 (51.0%) | 705 (50.4%) | 5278 (50.6%) |
| S24. The supervision I receive in my practices<br>fields is good | 1540 (37.8%) | 2495 (50.3%) | 846 (60.5%) | 4881 (46.8%) |
| S28. I am willing to take risks by participating in<br>practical in the context of pandemic | 904 (22.2%) | 2785 (56.1%) | 1141 (81.6%) | 4830 (46.3%) |
| S20. I prefer to study theoretical content using<br>distance learning methods | 1810 (44.4%) | 2181 (43.9%) | 566 (40.5%) | 4557 (43.7%) |
| S10. It is the duty of the medical student to put<br>himself at the service of the population in the<br>pandemic | 691 (17.0%) | 2525 (50.9%) | 1260 (90.1%) | 4476 (42.9%) |
| S25. I have access to psychological support | 1384 (34.0%) | 2212 (44.6%) | 783 (56.0%) | 4379 (42.0%) |
| S18. After pandemic, only practical academic<br>activities must be restored | 1658 (40.7%) | 2075 (41.8%) | 616 (44.1%) | 4349 (41.7%) |
| S21. My emotional state during the pandemic<br>affects my learning | 1939 (47.6%) | 1828 (36.8%) | 497 (35.6%) | 4264 (40.9%) |
| S7. I feel able to communicate the diagnosis for<br>COVID-19 infection | 1167 (28.7%) | 2062 (41.5%) | 782 (55.9%) | 4011 (38.4%) |
| S23. I feel stressed in the hospital at the moment | 1974 (48.5%) | 1371 (27.6%) | 264 (18.9%) | 3609 (34.6%) |
| S16. I prefer to delay my training to fully replace<br>academic activities than to participate in<br>distance learning activities | 1373 (33.7%) | 1602 (32.3%) | 544 (38.9%) | 3519 (33.7%) |
| S6. I am able to participate in the care of patients<br>who seek health care | 807 (19.8%) | 1891 (38.1%) | 751 (53.7%) | 3449 (33.1%) |
| S14. Medical schools must suspend their<br>academic activities during internship | 2236 (54.9%) | 832 (16.8%) | 298 (21.3%) | 3366 (32.3%) |
| S4. I know how to guide for therapeutic<br>measures | 1007 (24.7%) | 1689 (34.0%) | 564 (40.3%) | 3260 (31.2%) |
| S27. The role of the medical students during the<br>pandemics time is irrelevant | 444 (10.9%) | 144 (2.9%) | 76 (5.4%) | 664 (6.4%) |

Table 4 shows the adjusted odds ratios (and 95% confidence intervals), from multinomial models, for the association between students’ characteristics and perceptions and their view about the role of medical students during the COVID-19 pandemic (crude models results are presented in Supplemental Table 2). Compared to the participation of no medical students, odds ratios [OR] for the participation of students in internship or all medical students were, respectively, 0.61 (95% confidence interval: 0.55–0.67) and 0.13 (95% CI: 0.10–0.16). Men were more prone to support the participation of medical students in the fight against the pandemic (OR for the participation of students in internship or all medical students respectively 1.36 [95%CI: 1.24–1.49] and 1.68 [95%CI:1.47–1.91]).

**Table 4:** Adjusted odds ratios (95% confidence intervals) for the association between students’ characteristics and perceptions and their view about the role of medical students during the COVID-19 pandemic.

|  | No students should participate | Only students in internship should participate | All students should participate |
| --- | --- | --- | --- |
| **Year of medical school** |  |  |  |
| First/second (Basic sciences) | 1.0 (Reference) | Reference | Reference |
| Third/fourth (Clinical sciences) | 1.0 (Reference) | **0.72 (0.65 - 0.79)** | **0.60 (0.53 - 0.69)** |
| Fifth/sixth (Internship) | 1.0 (Reference) | **0.61 (0.55 - 0.67)** | **0.13 (0.10 - 0.16)** |
| **Sex** |  |  |  |
| Female | 1.0 (Reference) | 1.0 (Reference) | 1.0 (Reference) |
| Male | 1.0 (Reference) | **1.36 (1.24 - 1.49)** | **1.68 (1.47 - 1.91)** |
| **Personal / family / friend diagnosis of COVID-19** |  |  |  |
| No | 1.0 (Reference) | 1.0 (Reference) | 1.0 (Reference) |
| Yes | 1.0 (Reference) | 0.91 (0.77 - 1.07) | 0.87 (0.68 - 1.12) |
| **Beliefs towards a participation of medical students in COVID-19 pandemic healthcare** |  |  |  |
| S10. It is the duty of the medical student to put himself at the service of the population in the pandemic | 1.0 (Reference) | **5.03 (4.55 - 5.56)** | **44.10 (36.25 - 53.65)** |
| S28. I am willing to take risks by participating in practical in the context of pandemic | 1.0 (Reference) | **5.04 (4.57 - 5.55)** | **20.34 (17.32 - 23.90)** |
| S6. I am able to participate in the care of patients who seek health care | 1.0 (Reference) | **3.55 (3.18 - 3.96)** | **10.74 (9.23 - 12.50)** |
| S7. I feel able to communicate the diagnosis for COVID-19 infection | 1.0 (Reference) | **2.15 (1.96 - 2.37)** | **5.33 (4.63 - 6.12)** |
| S22. I will be a better health professional for having experienced the pandemic | 1.0 (Reference) | **2.13 (1.95 - 2.32)** | **3.56 (3.11 - 4.08)** |
| S4. I know how to guide for therapeutic measures | 1.0 (Reference) | **1.83 (1.66 - 2.02)** | **3.20 (2.79 - 3.67)** |
| S24. The supervision I receive in my practices fields is good | 1.0 (Reference) | **1.74 (1.60 - 1.89)** | **2.84 (2.50 - 3.23)** |
| S3. I know how to guide for preventive measures | 1.0 (Reference) | **2.39 (1.95 - 2.94)** | **2.33 (1.69 - 3.22)** |
| S1. I feel prepared to identify a suspected case | 1.0 (Reference) | **1.59 (1.45 - 1.74)** | **2.32 (2.02 - 2.65)** |
| S2. I can identify signs of severity in a patient | 1.0 (Reference) | **1.48 (1.34 - 1.64)** | **2.32 (1.99 - 2.70)** |
| S5. I know how to use personal protection equipment (PFE) | 1.0 (Reference) | **1.68 (1.52 - 1.86)** | **2.22 (1.87 - 2.62)** |
| S25. I have access to psychological support | 1.0 (Reference) | **1.47 (1.34 - 1.60)** | **2.12 (1.87 - 2.40)** |
| S17. After pandemic, academic activities must be fully restored | 1.0 (Reference) | 1.09 (1.00 - 1.18) | **1.39 (1.22 - 1.59)** |
| S26. I am proud of the way my institution responded to social and health demands in the face of the pandemic | 1.0 (Reference) | **1.29 (1.18 - 1.40)** | **1.36 (1.20 - 1.54)** |
| S18. After pandemic, only practical academic activities must be restored | 1.0 (Reference) | 1.07 (0.99 - 1.17) | **1.21 (1.07 - 1.38)** |
| S19. I feel able to study my medical course contents at distance | 1.0 (Reference) | **1.10 (1.01 - 1.20)** | **1.19 (1.05 - 1.35)** |
| S16. I prefer to delay my training to fully replace academic activities than to participate in distance learning activities | 1.0 (Reference) | **0.89 (0.82 - 0.98)** | 1.10 (0.97 - 1.26) |
| **Beliefs not / poorly related with the participation of medical students in COVID-19 pandemic healthcare** |  |  |  |
| S20. I prefer to study theoretical content using distance learning methods | 1.0 (Reference) | 1.05 (0.97 - 1.15) | 1.04 (0.91 - 1.18) |
| S15. Distance learning must be implemented in the suspension of academic activities | 1.0 (Reference) | 0.99 (0.91 - 1.08) | 1.05 (0.93 - 1.20) |
| **Beliefs against a participation of medical students in COVID-19 pandemic healthcare** |  |  |  |
| S14. Medical schools must suspend their academic activities during internship | 1.0 (Reference) | **0.17 (0.15 - 0.19)** | **0.26 (0.22 - 0.30)** |
| S13. Medical schools must suspend their academic activities during the first to fourth year. | 1.0 (Reference) | **0.73 (0.65 - 0.82)** | **0.26 (0.23 - 0.30)** |
| S23. I feel stressed in the hospital at the moment | 1.0 (Reference) | **0.43 (0.40 - 0.47)** | **0.31 (0.27 - 0.36)** |
| **S12. I am afraid of contaminating myself** | 1.0 (Reference) | **0.53 (0.49 - 0.58)** | **0.39 (0.34 - 0.44)** |
| **S27. The role of the medical students during the pandemics time is irrelevant** | 1.0 (Reference) | **0.26 (0.21 - 0.32)** | **0.60 (0.47 - 0.78)** |
| **S21. My emotional state during the pandemic affects my learning** | 1.0 (Reference) | **0.66 (0.61 - 0.72)** | **0.65 (0.57 - 0.74)** |
| **S11. I feel insecure regarding the future** | 1.0 (Reference) | **0.79 (0.72 - 0.86)** | **0.73 (0.64 - 0.83)** |

Beliefs towards a participation of medical students in COVID-19 pandemic healthcare with highest odds ratios were:

S10 “It is the duty of the medical student to put himself at the service of the population in the pandemic”,

S28 “I am willing to take risks by participating in practical in the context of pandemic”,

S6 “I am able to participate in the care of patients who seek health care”,

S7 “I feel able to communicate the diagnosis for COVID-19 infection”, and

S22 “I will be a better health professional for having experienced the pandemic”.

On the other hand, beliefs against a participation of medical students in COVID-19 pandemic healthcare with lowest odds ratios were statements S13/S14 (suspension of academic activities), S11 “I feel insecure regarding the future”, S21 “My emotional state during the pandemic affects my learning”, and

S27 “The role of the medical students during the pandemics time is irrelevant”.

Analysis with only the students in the last years (interns) was also performed (Table 5). In this subgroup, male sex was also associated with the support of a participation of medical students. Similar to the findings of the entire sample, statements S10, S28, S6, S2, S4 and S22 were the beliefs towards a participation of medical students with highest odds ratios in this subgroup. On the other hand, agreement with the statements S13/S14 (suspension of academic activities), S23 “I feel stressed in the hospital at the moment”, S12 “I am afraid of contaminating myself” and S27 “The role of the medical students during the pandemics time is irrelevant” were more relevant beliefs associated with a perception against the participation of medical students in the actions during the pandemic. Crude models for this subgroup are presented in Supplemental Table 3.

**Table 5:** Adjusted odds ratios (95% confidence intervals) for the association between internship students’ characteristics and perceptions and their view about the role of medical students during the COVID-19 pandemic.

|  | No students should participate | Only students in internship should participate | All students should participate |
| --- | --- | --- | --- |
| **Sex** |  |  |  |
| Female | 1.0 (Reference) | 1.0 (Reference) | 1.0 (Reference) |
| Male | 1.0 (Reference) | **1.29 (1.08 - 1.54)** | **1.75 (1.13 - 2.73)** |
| **Personal / family / friend diagnosis of COVID-19** |  |  |  |
| No | 1.0 (Reference) | 1.0 (Reference) | 1.0 (Reference) |
| Yes | 1.0 (Reference) | 0.79 (0.58 - 1.06) | 0.46 (0.17 - 1.29) |
| **Beliefs towards a participation of medical students in COVID-19 pandemic healthcare** |  |  |  |
| S10. It is the duty of the medical student to put himself at the service of the population in the pandemic | 1.0 (Reference) | **7.18 (5.86 - 8.79)** | **22.93 (13.45 - 39.10)** |
| S28. I am willing to take risks by participating in practical in the context of pandemic | 1.0 (Reference) | **12.24 (10.04 - 14.92)** | **13.26 (7.66 - 22.97)** |
| S6. I am able to participate in the care of patients who seek health care | 1.0 (Reference) | **6.86 (5.69 - 8.26)** | **6.29 (3.79 - 10.44)** |
| S2. I can identify signs of severity in a patient | 1.0 (Reference) | **3.31 (2.47 - 4.44)** | **5.70 (1.78 - 18.25)** |
| S4. I know how to guide for therapeutic measures | 1.0 (Reference) | **2.70 (2.28 - 3.20)** | **4.63 (2.84 - 7.57)** |
| S22. I will be a better health professional for having experienced the pandemic | 1.0 (Reference) | **3.76 (3.16 - 4.48)** | **4.46 (2.72 - 7.32)** |
| S24. The supervision I receive in my practices fields is good | 1.0 (Reference) | **2.75 (2.32 - 3.26)** | **4.25 (2.65 - 6.80)** |
| S7. I feel able to communicate the diagnosis for COVID-19 infection | 1.0 (Reference) | **3.30 (2.77 - 3.93)** | **4.06 (2.45 - 6.72)** |
| S25. I have access to psychological support | 1.0 (Reference) | **2.12 (1.77 - 2.55)** | **3.50 (2.26 - 5.43)** |
| S5. I know how to use personal protection equipment (PFE) | 1.0 (Reference) | **2.58 (2.10 - 3.15)** | **2.79 (1.53 - 5.10)** |
| S3. I know how to guide for preventive measures | 1.0 (Reference) | **5.40 (3.14 - 9.30)** | 2.27 (0.70 - 7.35) |
| S26. I am proud of the way my institution responded to social and health demands in the face of the pandemic | 1.0 (Reference) | 1.14 (0.97 - 1.35) | **1.73 (1.12 - 2.66)** |
| S19. I feel able to study my medical course contents at distance | 1.0 (Reference) | 1.01 (0.86 - 1.20) | **1.68 (1.05 - 2.67)** |
| S17. After pandemic, academic activities must be fully restored | 1.0 (Reference) | 0.87 (0.73 - 1.02) | **1.64 (1.05 - 2.56)** |
| S1. I feel prepared to identify a suspected case | 1.0 (Reference) | **2.67 (2.17 - 3.28)** | 1.63 (0.96 - 2.75) |
| **Beliefs not / poorly related with the participation of medical students in COVID-19 pandemic healthcare** |  |  |  |
| S20. I prefer to study theoretical content using distance learning methods | 1.0 (Reference) | 0.85 (0.72 - 1.00) | 1.01 (0.65 - 1.55) |
| S18. After pandemic, only practical academic activities must be restored | 1.0 (Reference) | 1.01 (0.86 - 1.20) | 1.40 (0.91 - 2.15) |
| **Beliefs against a participation of medical students in COVID-19 pandemic healthcare** |  |  |  |
| S13. Medical schools must suspend their academic activities during the first to fourth year. | 1.0 (Reference) | 0.79 (0.59 - 1.04) | **0.21 (0.13 - 0.34)** |
| S14. Medical schools must suspend their academic activities during internship | 1.0 (Reference) | **0.16 (0.14 - 0.20)** | **0.29 (0.19 - 0.46)** |
| S23. I feel stressed in the hospital at the moment | 1.0 (Reference) | **0.28 (0.23 - 0.33)** | **0.44 (0.28 - 0.68)** |
| S12. I am afraid of contaminating myself | 1.0 (Reference) | **0.37 (0.31 - 0.44)** | **0.53 (0.33 - 0.84)** |
| S27. The role of the medical students during the pandemics time is irrelevant | 1.0 (Reference) | **0.14 (0.10 - 0.20)** | 0.65 (0.35 - 1.22) |
| **S21. My emotional state during the pandemic affects my learning** | 1.0 (Reference) | **0.49 (0.41 - 0.58)** | 0.74 (0.47 - 1.14) |
| **S16. I prefer to delay my training to fully replace academic activities than to participate in distance learning activities** | 1.0 (Reference) | **0.69 (0.58 - 0.83)** | 1.00 (0.63 - 1.58) |
| **S15. Distance learning must be implemented in the suspension of academic activities** | 1.0 (Reference) | **0.81 (0.68 - 0.96)** | 1.03 (0.65 - 1.65) |
| **S11. I feel insecure regarding the future** | 1.0 (Reference) | **0.69 (0.58 - 0.83)** | 1.06 (0.64 - 1.76) |

## Discussion

In this study we aimed to evaluate the factors that motivate a medical student to participate in the care of patients in the context of COVID-19 pandemic. We also aimed to understand the students’ desires related to continuing medical training or to have online education during the pandemic. To our knowledge, there is no previous study that evaluated a large sample of medical students with these purposes. We believe that this study can contribute to the planning of medical education and work force organization in the COVID-19 pandemic and in other future health emergencies. Although our sample was a convenience sample, we evaluated a large group of medical students (10,433) and there was a uniform distribution of subjects across the country (Table 1).

When we performed this study, the number of COVID-19 cases in Brazil was low, and only 7·0% of participants (Table 3) had a family member or a friend with a diagnosis of COVID-19 or had a personal diagnosis of this disease. [10] We were able to study the factors that influence the desire to participate in the care of COVID-19 without a strong emotional influence of sick relatives or friends.

When we compared the answers to the statements of the questionnaire provided by students in the first two years of medical course to students of the last two years (interns) (Table 2, univariate analysis) we observed many differences.

The largest differences in percentages of agreement with the statements were related to feeling of competency to take care of patients (differences from 26.7% to 37.7%, statements S1, S2, S4, S6 and S7). As expected, interns felt more confident concerning identification of a suspected case, identification of signs of severity, guidance of therapeutic measures, participation in the care of patients and communication of diagnosis. In addition, interns were more prone to accept on-line learning and the interruption of both classes and internship activities (differences from 16.0% to 19.0%). In contrast, students from the first years were more insecure about the substitution of classes and practical activities by on-line learning (Statements S16 and S17, differences 10.5% and 17.9%). More students preferred to delay their training and fully restore the academic activities.

Although interns felt more secure concerning competencies to take care of COVID-19 patients, many interns probably needed psychological support. In fact, more interns agreed to the statement S23 (“I feel stressed in the hospital at the moment”, difference 24.5% compared to first year medical students) and less interns agreed with statement S25 (“I have access to psychological support”, difference 17.6%). In addition, many students were afraid of contaminating themselves (Statement S12, percentages of agreement from 59.2 to 68.4%) and considered that their emotional state affected learning (Statement S21, percentages of agreement from 40.6 to 44.5%).

To perform the multivariate analysis, we decided to divide the medical students into three groups, concerning their responses about participation of medical students in the COVID-19 pandemic (no participation, only interns participate or all students participate, Table 3), to better understand the factors that were most important in the student’s decision to participate. Since we observed many differences between the answers of students in the first years of medical program and interns, we performed two analysis, one including all medical students and the second including only interns (Tables 4 and 5). Interestingly, we did not observe important differences when we compared the two analysis, suggesting that individual factors are more important than professional identity developed during medical course. Maybe the decision to be a volunteer in a health emergency such as COVID-19 pandemic is more linked to an emotional or attitudinal decision than beliefs of self-efficacy or performance.

The statements that had greater odds ratios when the groups of “all students should participate” and “only students in internship should participate” were compared to “no students should participate” group (reference, odds 1.0) were, for all students, statements S10, S28, S6, S7 and S22, and for interns only, statements S10, S28, S6, S2, S4 and S22.

We observed that sense of purpose or duty (moral values linked to medical profession) (S10 “It is the duty of the medical student to put himself at the service of the population in the pandemic”) was the most important factor that influenced the desire to work in the pandemic, followed by the willingness to take risk (altruism) (S28 “I am willing to take risks by participating in practical in the context of pandemic”) and the perception of good performance (self-efficacy) (S6 “I am able to participate in the care of patients who seek health care” and S7 “I feel able to communicate the diagnosis for COVID-19 infection”, for all medical students and S6, S2 “I can identify signs of severity in a patient” and S4 “I know how to guide for therapeutic measures”) for interns. In addition to statements that suggest moral values, altruism and confidence in professional competence, in both analysis, statement S22 (“I will be a better health professional for having experienced the pandemic”), that is related to building of professional identity, had odds ratios above 2.0 in all comparisons. In other words, the desire to help was stronger among the students that believed that they should work in the pandemic than their interest in learning during this health emergency. Allowing students to participate can reinforce important values, such as altruism, service in times of crisis, and solidarity with the profession, contributing to the building of professional identity. [12]

Why people act or decide to serve others is important to better organize volunteerism in a pandemic. Clary et al. studied volunteers of different areas and observed six different motivation functions to be a volunteer: value (opportunities to express altruism and humanitarian values), understanding (opportunities to learn something new or to develop skills), social (opportunities to stablish relationships), career (opportunities to career benefits), protective (scape from their negative feelings) and enhancement (opportunities to add self-esteem). [13] Guntert et al., studying 2,017 volunteers, divided these functions in two categories self-determined and controlled motivation. They included in the self-determined category the functions altruism and humanitarian values and understanding motives, and in the controlled motivation the other functions as enhancement, protective, social and career. [14] They concluded that self-determined motivation results in more volunteer’s satisfaction. [14]

In our study we observed that the student with higher sense of duty (S10) and altruism (S28) were more prone to engage in health care activities during COVID-19, and it could be interpreted as self-determined motivation or values function of motivation. We also observed that the self-perception of competence (S6 and S7) was the third factor influencing motivation and the fourth was the desire of learn with the experience of working in the pandemic. Desire to learn demonstrated for student can be interpreted as understanding function by Clary et al., and it is also a self-determined motivation defined by Guntert et al. [13,14] The controlled motivation was not important in our data.

We also evaluated factors that can influence the decision to not participate in the COVID-19 health effort. We observed that predictors of do not be a volunteer were beliefs that all educational activities should be suspended (S13 and S14), fear of contamination (S12 “I am afraid of contaminating myself”) and emotional factors (S23 “I feel stressed in the hospital at the moment”).

The differences we observed between male and female medical students were smaller than the differences among students of different years of medical program (Table 2, univariate analysis). Differences between males and females concerning the percentage of agreement with the statements of the questionnaire were greater than 9% in only 5 statements (females vs males): S11 “I feel insecure regarding the future” (16.0%), S7 “I feel able to communicate the diagnosis for COVID-19 infection” (−13.8%), S21 “My emotional state during the pandemic affects my learning” (11.2%), S28 “I am willing to take risks by participating in practical activities in the context of pandemic” (−10.0%) and S12 “I am afraid of contaminating myself” (9.7%). All these statements are related to emotional competencies. Results from multinomial regressions performed with all medical students (Table 3) and with only interns (Table 4) also showed that males were more prone to believe that only interns should participate (odds rations 1.36 and 1.29, respectively, for all students and interns) and that all students should participate (odds ratios 1.68 and 1.75, respectively). We argue that this difference is possibly due males’ higher propensity to take risks in health/safety domain. [15] Also, females are more prone to develop anxiety and stress disorders, be more affected by human suffering and have worse perceptions about their own quality of life, health and skills. [16–18] These factors can influence their intentions to act in pandemic. Nonetheless, we cannot disregard gender bias as a possible limitation of our instrument. [18,19]

COVID-19 pandemic had a substantial impact on medical education across the world. [3,5] There is uncertainty and disagreement about the appropriate roles for medical students during this pandemic, and student participation in clinical care has varied across institutions and countries. [4,12] Many medical schools forbid any patient interaction, others included medical students in patient care and others decided to graduate medical students early in order to make them frontline clinicians. The American Association of Medical Colleges recommended that “unless there is a critical health care workforce need locally, we strongly suggest that medical students not be involved in any direct patient care activities”. [4] However, some medical educators have a different point of view, considering that medical schools should offer students clinical opportunities that would benefit patient care and potentially help to prevent workforce shortages. [20,21] These different attitudes may have implications for medical education concerning the role of medical students in a local or global health emergency. Some educators chose the position to ensure student security and to avoid exposition, and this perspective can send a message (hidden curriculum) that students have a passive role without a significant social responsibility. On the other hand, medical educators that defend the inclusion of the medical student in health teams send to medical students a message that social responsibility is pivotal to professional identity. Interestingly, in our study, 56.9% of medical students agreed with statement S22 (“I will be a better health professional for having experienced the pandemic”).

A shortage of health professionals has been reached even in some cities of developed countries. [20,21] This risk will be even greater as the pandemic is reaching developing countries in South America, Asia and Africa. The participation of medical students in the care of people with suspected or confirmed COVID-19 increases personal risk of acquiring this disease. However, the risks of severe disease are probably lower than to retired clinician volunteers, more susceptible to complications of COVID-19 owing to their age. [22] Since the personal risks cannot be eliminated, there is predominant agreement that involvement of medical students in the care of patients should be voluntary. [5,6] Medical students that work as volunteers, must have appropriate training, do not undertake any activity beyond their level of competence, have continuous supervision and adequate personal protective equipment. [5,6]

One implication of our study for medical education is that allowing students to participate in pandemic efforts reinforces important values, such as altruism, service in times of crisis, and solidarity with the profession and disposition to serve society. [12] It will probably influence the development of professional values and identity.

## Conclusions

Our study showed that medical students who believe that they must participate in the fight against COVID-19 pandemic are motivated by sense of purpose or duty, altruism, perception of good performance and values of professionalism more than their interest in learning. These results have implications in the developing of programs of volunteering and in the design of health force policies in the present pandemic and in future health emergencies.

## Data Availability

All data is available upon request to the corresponding author.

## Authors’ contributions

Study design and data collection: P Tempski, FM Arantes-Costa, MB Torsani, MAM

Siqueira, BQRC Amaro, MEFM Nascimento, SL Siqueira and MA Martins

Literature search: FM Arantes-Costa, MAM Siqueira and MA Martins

Data analysis and data interpretation: P Tempski, R Kobayasi, IS Santos and MA Martins

Revising the manuscript: all authors

Final approval of the version to be published: all authors

Agreement to be accountable for all aspects of the work: all authors

## Acknowledgement

The authors wish to thank the Brazilian Section of the International Federation of Medical Students Associations (IFMSA) for the help during data collection of the study.

## Supplemental Tables

**Supplemental Table 1:** Distribution of answers by each statement, all participants (n = 10,433).

|  | Agree | Not agree or disagree | Disagree |
| --- | --- | --- | --- |
| S1 I feel prepared to identify a suspected case | 6393 (61.3) | 2117 (20.3) | 1923 (18.4) |
| S2 I can identify signs of severity in a patient | 7650 (73.3) | 1504 (14.4) | 1279 (12.3) |
| S3 I know how to guide for preventive measures | 9967 (95.5) | 317 (3.0) | 149 (1.4) |
| S4 I know how to guide for therapeutic measures | 3260 (31.2) | 2862 (27.2) | 4311 (41.3) |
| S5 I know how to use personal protection equipment (PFE) | 8255 (79.1) | 1366 (13.1) | 812 (7.8) |
| S6 I am able to participate in the care of patients who seek health care | 3449 (33.1) | 2757 (26.4) | 4227 (40.5) |
| S7 I feel able to communicate the diagnosis for COVID-19 infection | 4001 (38.4) | 2373 (22.7) | 4049 (38.8) |
| S8 Medical internship students must participate in health care assistance during pandemic | 6290 (60.3) | 2619 (25.1) | 1524 (14.6) |
| S9 All students regardless their year during the course, must participate in health care assistance during pandemic | 1398 (13.4) | 2132 (20.4) | 6903 (66.2) |
| S10 It is the duty of the medical student to put himself at the service of the population in the pandemic | 4460 (42.9) | 3073 (29.5) | 2884 (27.6) |
| S11 I feel insecure regarding the future | 6769 (64.9) | 1870 (17.9) | 1794 (17.2) |
| S12 I am afraid of contaminating myself | 6433 (61.7) | 2020 (19.4) | 1980 (19.0) |
| S13 Medical schools must suspend their academic activities during the first to fourth year. | 8125 (77.9) | 1393 (13.4) | 915 (8.8) |
| S14 Medical schools must suspend their academic activities during internship | 3366 (32.3) | 3243 (31.1) | 3824 (36.7) |
| S15 Distance learning must be implemented in the suspension of academic activities | 6142 (58.9) | 1982 (19.0) | 2309 (22.1) |
| S16 I prefer to delay my training to fully replace academic activities than to participate in distance learning activities | 3519 (33.7) | 2008 (19.2) | 4906 (47.0) |
| S17 After pandemic, academic activities must be fully restored | 6112 (58.6) | 2280 (21.9) | 2041 (19.6) |
| S18 After pandemic, only practical academic activities must be restored | 4349 (41.7) | 2052 (19.7) | 4032 (38.6) |
| S19 I feel able to study my medical course contents at distance | 5278 (50.6) | 2292 (22.0) | 2863 (27.4) |
| S20 I prefer to study theoretical content using distance learning methods | 4557 (43.7) | 2304 (22.1) | 3572 (34.2) |
| S21 My emotional state during the pandemic affects my learning | 4264 (40.9) | 2269 (21.7) | 3900 (37.4) |
| S22 I will be a better health professional for having experienced the pandemic | 5934 (56.9) | 3124 (29.9) | 1375 (13.2) |
| S23 I feel stressed in the hospital at the moment | 3609 (34.6) | 3717 (35.6) | 3107 (29.8) |
| S24 The supervision I receive in my practices fields is good | 4811 (46.8) | 3956 (37.9) | 1596 (15.3) |
| S25 I have access to psychological support | 4379 (42.0) | 2192 (21.0) | 3862 (37.0) |
| S26 I am proud of the way my institution responded to social and health demands in the face of the pandemic | 5549 (53.2) | 2781 (26.7) | 2103 (20.2) |
| S27 The role of the medical students during the pandemics time is irrelevant | 664 (6.4) | 1951 (18.7) | 7818 (74.9) |
| S28 I am willing to take risks by participating in practical in the context of pandemic | 4830 (46.3) | 2352 (22.5) | 3251 (31.2) |

**Supplemental Table 2:**
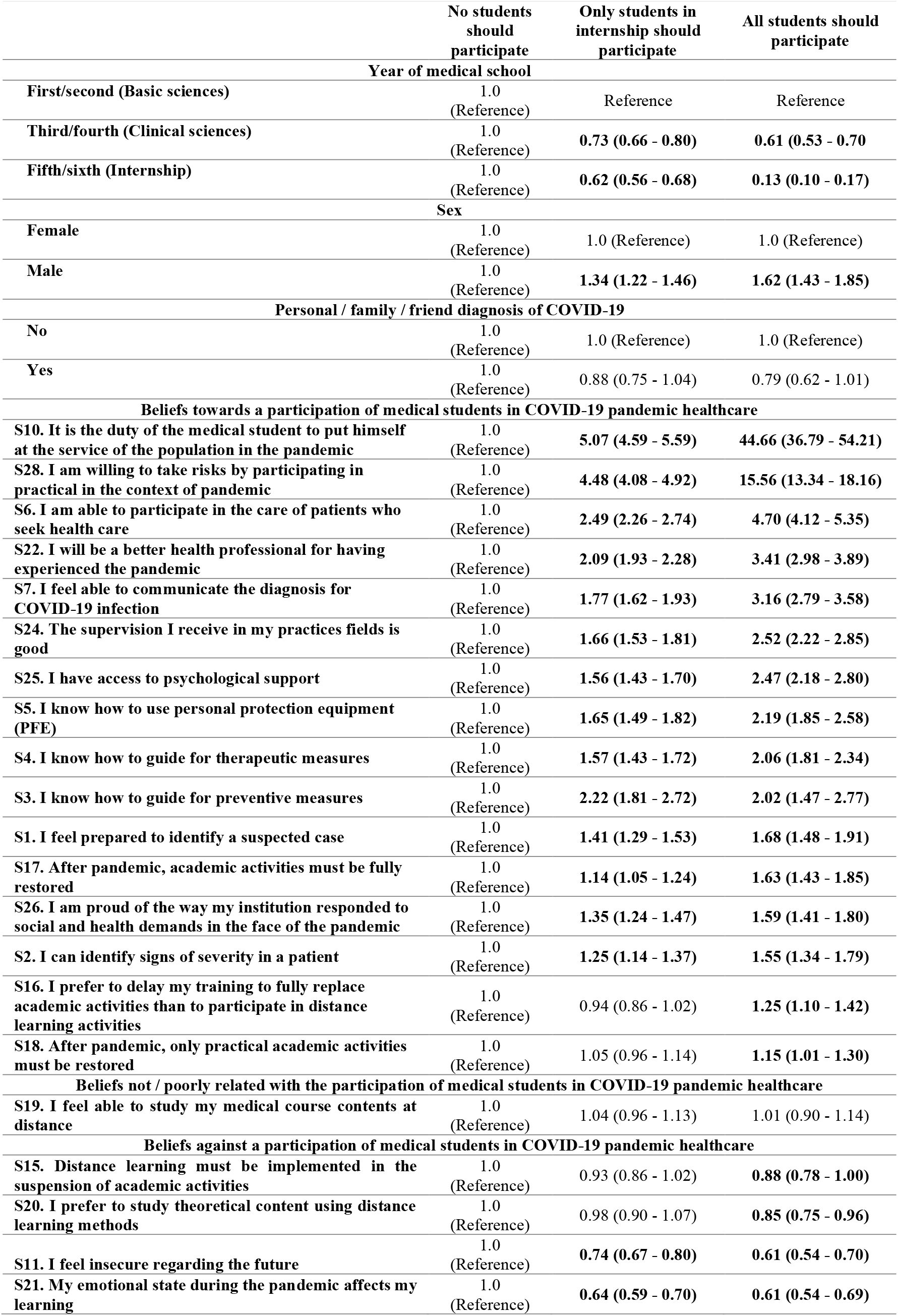
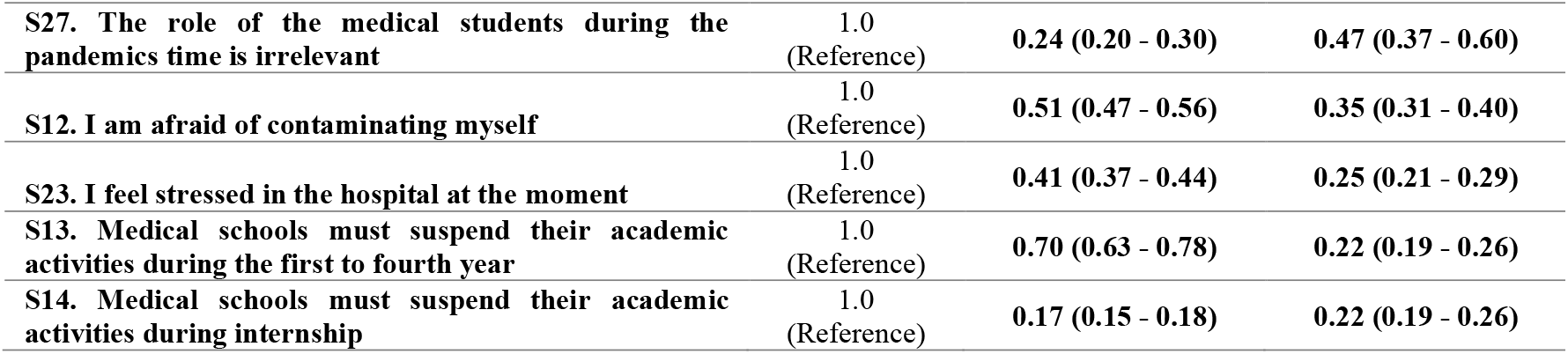
Crude odds ratios (95% confidence intervals) for the association between students’ characteristics and perceptions and their view about the role of medical students during the COVID-19 pandemic

|  | No students should participate | Only students in internship should participate | All students should participate |
| --- | --- | --- | --- |
| **Year of medical school** |  |  |  |
| First/second (Basic sciences) | 1.0<br>(Reference) | Reference | Reference |
| Third/fourth (Clinical sciences) | 1.0<br>(Reference) | **0.73 (0.66 - 0.80)** | **0.61 (0.53 - 0.70)** |
| Fifth/sixth (Internship) | 1.0<br>(Reference) | **0.62 (0.56 - 0.68)** | **0.13 (0.10 - 0.17)** |
| **Sex** |  |  |  |
| Female | 1.0<br>(Reference) | 1.0 (Reference) | 1.0 (Reference) |
| Male | 1.0<br>(Reference) | **1.34 (1.22 - 1.46)** | **1.62 (1.43 - 1.85)** |
| **Personal / family / friend diagnosis of COVID-19** |  |  |  |
| No | 1.0<br>(Reference) | 1.0 (Reference) | 1.0 (Reference) |
| Yes | 1.0<br>(Reference) | 0.88 (0.75 - 1.04) | 0.79 (0.62 - 1.01) |
| **Beliefs towards a participation of medical students in COVID-19 pandemic healthcare** |  |  |  |
| S10. It is the duty of the medical student to put himself at the service of the population in the pandemic | 1.0<br>(Reference) | **5.07 (4.59 - 5.59)** | **44.66 (36.79 - 54.21)** |
| S28. I am willing to take risks by participating in practical in the context of pandemic | 1.0<br>(Reference) | **4.48 (4.08 - 4.92)** | **15.56 (13.34 - 18.16)** |
| S6. I am able to participate in the care of patients who seek health care | 1.0<br>(Reference) | **2.49 (2.26 - 2.74)** | **4.70 (4.12 - 5.35)** |
| S22. I will be a better health professional for having experienced the pandemic | 1.0<br>(Reference) | **2.09 (1.93 - 2.28)** | **3.41 (2.98 - 3.89)** |
| S7. I feel able to communicate the diagnosis for COVID-19 infection | 1.0<br>(Reference) | **1.77 (1.62 - 1.93)** | **3.16 (2.79 - 3.58)** |
| S24. The supervision I receive in my practices fields is good | 1.0<br>(Reference) | **1.66 (1.53 - 1.81)** | **2.52 (2.22 - 2.85)** |
| S25. I have access to psychological support | 1.0<br>(Reference) | **1.56 (1.43 - 1.70)** | **2.47 (2.18 - 2.80)** |
| S5. I know how to use personal protection equipment (PFE) | 1.0<br>(Reference) | **1.65 (1.49 - 1.82)** | **2.19 (1.85 - 2.58)** |
| S4. I know how to guide for therapeutic measures | 1.0<br>(Reference) | **1.57 (1.43 - 1.72)** | **2.06 (1.81 - 2.34)** |
| S3. I know how to guide for preventive measures | 1.0<br>(Reference) | **2.22 (1.81 - 2.72)** | **2.02 (1.47 - 2.77)** |
| S1. I feel prepared to identify a suspected case | 1.0<br>(Reference) | **1.41 (1.29 - 1.53)** | **1.68 (1.48 - 1.91)** |
| S17. After pandemic, academic activities must be fully restored | 1.0<br>(Reference) | **1.14 (1.05 - 1.24)** | **1.63 (1.43 - 1.85)** |
| S26. I am proud of the way my institution responded to social and health demands in the face of the pandemic | 1.0<br>(Reference) | **1.35 (1.24 - 1.47)** | **1.59 (1.41 - 1.80)** |
| S2. I can identify signs of severity in a patient | 1.0<br>(Reference) | **1.25 (1.14 - 1.37)** | **1.55 (1.34 - 1.79)** |
| S16. I prefer to delay my training to fully replace academic activities than to participate in distance learning activities | 1.0<br>(Reference) | 0.94 (0.86 - 1.02) | **1.25 (1.10 - 1.42)** |
| S18. After pandemic, only practical academic activities must be restored | 1.0<br>(Reference) | 1.05 (0.96 - 1.14) | **1.15 (1.01 - 1.30)** |
| **Beliefs not / poorly related with the participation of medical students in COVID-19 pandemic healthcare** |  |  |  |
| S19. I feel able to study my medical course contents at distance | 1.0<br>(Reference) | 1.04 (0.96 - 1.13) | 1.01 (0.90 - 1.14) |
| **Beliefs against a participation of medical students in COVID-19 pandemic healthcare** |  |  |  |
| S15. Distance learning must be implemented in the suspension of academic activities | 1.0<br>(Reference) | 0.93 (0.86 - 1.02) | **0.88 (0.78 - 1.00)** |
| S20. I prefer to study theoretical content using distance learning methods | 1.0<br>(Reference) | 0.98 (0.90 - 1.07) | **0.85 (0.75 - 0.96)** |
| S11. I feel insecure regarding the future | 1.0<br>(Reference) | **0.74 (0.67 - 0.80)** | **0.61 (0.54 - 0.70)** |
| S21. My emotional state during the pandemic affects my learning | 1.0<br>(Reference) | **0.64 (0.59 - 0.70)** | **0.61 (0.54 - 0.69)** |
| **S27. The role of the medical students during the pandemics time is irrelevant** | 1.0<br>(Reference) | **0.24 (0.20 - 0.30)** | **0.47 (0.37 - 0.60)** |
| **S12. I am afraid of contaminating myself** | 1.0<br>(Reference) | **0.51 (0.47 - 0.56)** | **0.35 (0.31 - 0.40)** |
| **S23. I feel stressed in the hospital at the moment** | 1.0<br>(Reference) | **0.41 (0.37 - 0.44)** | **0.25 (0.21 - 0.29)** |
| **S13. Medical schools must suspend their academic activities during the first to fourth year** | 1.0<br>(Reference) | **0.70 (0.63 - 0.78)** | **0.22 (0.19 - 0.26)** |
| **S14. Medical schools must suspend their academic activities during internship** | 1.0<br>(Reference) | **0.17 (0.15 - 0.18)** | **0.22 (0.19 - 0.26)** |

**Supplemental Table 3:** Crude odds ratios (95% confidence intervals) for the association between internship students’ characteristics and perceptions and their view about the role of medical students during the COVID-19 pandemic.

|  | No students should participate | Only students in internship should participate | All students should participate |
| --- | --- | --- | --- |
| **Sex** |  |  |  |
| Female | 1.0 (Reference) | 1.0 (Reference) | 1.0 (Reference) |
| Male | 1.0 (Reference) | 1.22 (1.03 - 1.46) | 1.70 (1.09 - 2.63) |
| **Personal / family / friend diagnosis of COVID-19** |  |  |  |
| No | 1.0 (Reference) | 1.0 (Reference) | 1.0 (Reference) |
| Yes | 1.0 (Reference) | 0.79 (0.58 - 1.06) | 0.46 (0.17 - 1.29) |
| **Beliefs towards a participation of medical students in COVID-19 pandemic healthcare** |  |  |  |
| S10. It is the duty of the medical student to put himself at the service of the population in the pandemic | 1.0 (Reference) | 7.15 (5.85 - 8.74) | 23.16 (13.60 - 39.41) |
| S28. I am willing to take risks by participating in practical in the context of pandemic | 1.0 (Reference) | 12.45 (10.24 - 15.14) | 13.42 (7.79 - 23.13) |
| S6. I am able to participate in the care of patients who seek health care | 1.0 (Reference) | 6.99 (5.82 - 8.41) | 6.48 (3.92 - 10.72) |
| S2. I can identify signs of severity in a patient | 1.0 (Reference) | 3.31 (2.48 - 4.41) | 5.96 (1.87 - 19.01) |
| S4. I know how to guide for therapeutic measures | 1.0 (Reference) | 2.71 (2.29 - 3.20) | 4.78 (2.93 - 7.78) |
| S22. I will be a better health professional for having experienced the pandemic | 1.0 (Reference) | 3.79 (3.19 - 4.50) | 4.46 (2.72 - 7.31) |
| S24. The supervision I receive in my practices fields is good | 1.0 (Reference) | 2.88 (2.44 - 3.41) | 4.30 (2.70 - 6.85) |
| S7. I feel able to communicate the diagnosis for COVID-19 infection | 1.0 (Reference) | 3.36 (2.83 - 3.99) | 4.25 (2.58 - 7.03) |
| S25. I have access to psychological support | 1.0 (Reference) | 2.21 (1.85 - 2.65) | 3.57 (2.31 - 5.52) |
| S5. I know how to use personal protection equipment (PFE) | 1.0 (Reference) | 2.48 (2.03 - 3.03) | 2.76 (1.51 - 5.02) |
| S3. I know how to guide for preventive measures | 1.0 (Reference) | 5.34 (3.11 - 9.17) | 2.19 (0.68 - 7.08) |
| S26. I am proud of the way my institution responded to social and health demands in the face of the pandemic | 1.0 (Reference) | 1.14 (0.97 - 1.35) | 1.74 (1.13 - 2.68) |
| S1. I feel prepared to identify a suspected case | 1.0 (Reference) | 2.72 (2.21 - 3.33) | 1.72 (1.02 - 2.89) |
| S19. I feel able to study my medical course contents at distance | 1.0 (Reference) | 1.05 (0.89 - 1.24) | 1.70 (1.07 - 2.71) |
| **Beliefs not / poorly related with the participation of medical students in COVID-19 pandemic healthcare** |  |  |  |
| S17. After pandemic, academic activities must be fully restored | 1.0 (Reference) | 0.84 (0.71 - 0.98) | 1.66 (1.07 - 2.58) |
| S18. After pandemic, only practical academic activities must be restored | 1.0 (Reference) | 1.04 (0.88 - 1.23) | 1.40 (0.91 - 2.15) |
| S20. I prefer to study theoretical content using distance learning methods | 1.0 (Reference) | 0.86 (0.73 - 1.01) | 0.99 (0.64 - 1.52) |
| **Beliefs against a participation of medical students in COVID-19 pandemic healthcare** |  |  |  |
| S13. Medical schools must suspend their academic activities during the first to fourth year. | 1.0 (Reference) | 0.78 (0.59 - 1.03) | 0.21 (0.13 - 0.35) |
| S14. Medical schools must suspend their academic activities during internship | 1.0 (Reference) | 0.16 (0.13 - 0.19) | 0.29 (0.19 - 0.46) |
| S23. I feel stressed in the hospital at the moment | 1.0 (Reference) | 0.27 (0.22 - 0.32) | 0.41 (0.27 - 0.64) |
| S12. I am afraid of contaminating myself | 1.0 (Reference) | 0.35 (0.29 - 0.42) | 0.50 (0.32 - 0.79) |
| S27. The role of the medical students during the pandemics time is irrelevant | 1.0 (Reference) | 0.14 (0.10 - 0.20) | 0.67 (0.36 - 1.24) |
| S21. My emotional state during the pandemic affects my learning | 1.0 (Reference) | 0.47 (0.39 - 0.55) | 0.69 (0.45 - 1.07) |
| S11. I feel insecure regarding the future | 1.0 (Reference) | 0.68 (0.56 - 0.81) | 0.98 (0.60 - 1.61) |
| S15. Distance learning must be implemented in the suspension of academic activities | 1.0 (Reference) | 0.83 (0.70 - 0.98) | 1.00 (0.63 - 1.59) |
| S16. I prefer to delay my training to fully replace academic activities than to participate in distance learning activities | 1.0 (Reference) | 0.68 (0.56 - 0.81) | 1.02 (0.64 - 1.60) |

